# ‘Biohacking’: A thematic analysis of tweets to better understand how ‘biohackers’ conceptualise their practices

**DOI:** 10.1101/2023.02.16.23286022

**Authors:** Qasim Rafiq, Lynsey Christie, Heather May Morgan

## Abstract

Biohacking, considered to include technology such as wearables, lifestyle changes and nutrition to allow one to optimise their health, is growing in popularity. However, the definition of and insights according to those involved in these practices remains elusive and unexplored. Technological advancements, including the internet, have given rise to globally connected communities and various health-related consumer technologies that measure health metrics from the comfort of one’s own home. While health-related information sharing and technology-assisted health tracking may appear beneficial, it also affords many opportunities for harm through the spread of misinformation and the use of potentially inaccurate devices. Adopting a qualitative approach using thematic analysis, this study focused on identifying the practices and topics discussed publicly on the Twitter social media associated with the hashtags #biohacking and #biohacker. The main topics were physical fitness, nutrition, mental health, self-development, genetics, and neuroscience. Most of the biohacking practices were found to be health-centric and include practices such as dietary or herbal supplements or chip implants that could interact with medical investigations and treatments. This highlights that biohacking practices should be included as part of a proper medical history to allow healthcare providers to recommend safe and appropriate therapies, and to avoid supplement-drug interactions and adverse events. Implications for biohacking are vast and minimising harms, whilst optimising benefits at the individual and population level requires a better understanding of how biohacking practices are conceptualised. This will help inform healthcare decision-makers, policymakers, and industries associated with the practices identified.

**Author Summary:** Biohacking is growing in popularity and there is no published literature exploring exactly what this relatively new phenomenon entails and how biohackers conceptualise it. Published literature on biohacking often refers to the practise as involving invasive subdermal chip plants or various forms of technology. We searched Twitter using hashtags #biohacker and #biohacking to identify public tweets discussing this practise. We found the phenomenon of biohacking to be amorphous, encompassing a wide range of lifestyle measures, some of which do not require the use of technology such as nutrition and exercise. The advent of internet and technology has made health-related information sharing easily accessible across the globe, allowing users to track their health metrics and make changes without the input of a trained health professional. Implications for this are vast and includes many potential benefits but also many potential harms due to the spread of misinformation, and interactions between drugs and medical treatments. Our study provides new insights into how the emerging biohacking movement is conceptualised by biohackers on social media, implications for health safety and the need for a refined definition of biohacking to assist medical practitioners in talking with patients about their practises.

## Introduction

A dictionary definition of biohacking is: “attempts to improve the condition of your body and mind using technology, drugs, or other chemical substances such as hormones” [1]. This definition may be interpreted as attempts to improve health using technology, drugs, or other chemical substances such as hormones. Biohacking is growing in popularity; however, the definition of and insight from those involved in this practice remains elusive. Recent and more rapid advances in technological developments in the health domain, since the advent of the internet and emergence of personal computing devices, e.g., smartphones and wearables, have allowed innovation of a range of digital options and devices for health monitoring and information sharing [2]. Society has come a long way from some of the oldest known wearable technologies, e.g., the 17th-century Abacus ring originating from China’s Qing dynasty era to modern-day smartwatches and activity trackers [3], as show in in Figure 1.

**Figure 1:**
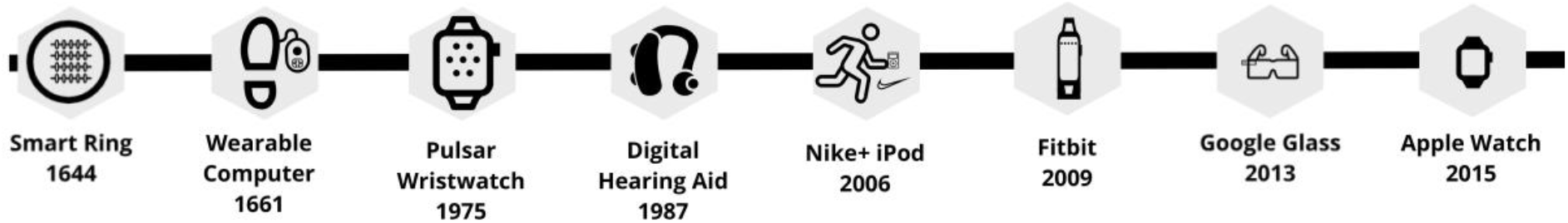
timeline of consumer wearables, adapted (The timeline of wearables. n.d.)

This includes both consumer grade wearables, e.g., fitness trackers such as Fitbit and Garmin, and medical-grade diagnostic devices e.g., Kardia, a pocket-sized electrocardiogram monitoring device [4,5]. Advances in these technologies combined with the internet boom has given rise to a health-conscious global movement of early adopters of healthcare technologies. Connected by networks of publicly shared tools and resources, an estimated 65.6% of the population were internet users in 2021, compared to 0.4% in 1995 [6,7]. This steep increase has been paralleled in the number of connected wearable devices bought/worn worldwide: sales more than doubled from 325 million in 2016 to 722 million in 2019, forecast to exceed 1 billion by 2022 [8]. This increase in internet users has facilitated sharing of health-related information with people wanting to change their lifestyle, such as nutrition and exercise habits based on this information, which may come from unreliable sources, potentially resulting in harm [9]. These related developments have sparked interest in the acquisition of and engagement with health data and do-it-yourself (DIY) biological modification, often undertaken in informal settings such as homemade laboratories [10], with knowledge and expertise coming from self-identifying biohacker communities, whether in person or online platforms [7], rather than in a formal medical context.

Technological advancements have also improved our understanding of nutrition, drugs and other chemicals, and this information is now disseminated and made accessible on blogs, social media, podcasts, and other online platforms [11,12]. Our understanding of the pharmacological effects of drugs and chemicals, conventionally used for prevention and management of medical conditions, has allowed for these to also be considered for use in health optimisation [12,13].

As adoption of unregulated consumer health-related technologies only continues to increase, it is imperative that healthcare staff are aware of the technological advancements occurring out with medical contexts. Awareness and adoption of healthcare related technologies into the healthcare system often lag behind the rapid progress of advancements due to strict safety regulations [14]. The risk of the public turning to biohacking practices to manage their health is likely to increase, requiring the crucial task of filling the gaps in our knowledge. Given the extensive nature of biohacking practices, it would be prudent for health care professionals to understand the fundamentals of biohacking to enable them to identify potential harms and mitigate these.

To the best of our knowledge, no published data exists about online discussions on biohacking, making it difficult to ascertain what exactly biohacking refers to in the eyes of the biohacking and non-biohacking public. There are a plethora of podcasts, blogs, websites, and conferences on biohacking, yet again there is ambiguity as to what exactly this refers to. The aim of this study was to explore what participants on Twitter (twitter.com) are discussing using the term ‘biohacking’, collecting tweets that included relevant hashtags (#biohacking and #biohacker) to identify the themes and topics discussed. This is the first known study to synthesise what biohacking practices entail and evaluate discussion on ‘biohacking’ on social media. This study attempts to investigate how biohackers conceptualise their practices and any associated technology to help fill this gap.

## Methods

### Data collection

Twitter is a social media platform that was established in 2006, allowing users to interact with each other, using 280-character messages, known as ‘tweets’ [15,16]. The public nature of tweets and the ease of accessing and searching the publicly available tweets makes Twitter ideal for mining data for research purposes. A hashtag used before a word or unbroken phrase creates a way to categorise tweets based on the content or target group of the tweet [15]. More than 300 million active users publish more than 500 million tweets daily, and whilst not representative of any specific population, the demographic on Twitter is broad [15].

Global tweets that included the English-language hashtags #biohacking or #biohacker were prospectively collected using Twitter archiving google spreadsheet (TAGS v6.1) between 24 August and 24 September 2020 [15,17]. These hashtags were selected to limit the analysis to tweets that users had identified as being relevant to biohacking. Only tweets including at least one of these hashtags will have been collected by TAGS for further analysis. TAGS allowed automated data retrieval over this period, using convenience sampling Tweets were collected and analysed by QR, excluding those that were irrelevant to the topic (promotions and spam not lending itself to meaningful thematic analysis) and those not in the English-language (see Figure 2). Independent coding of 10% of tweets was undertaken by HMM.

**Figure 2:**
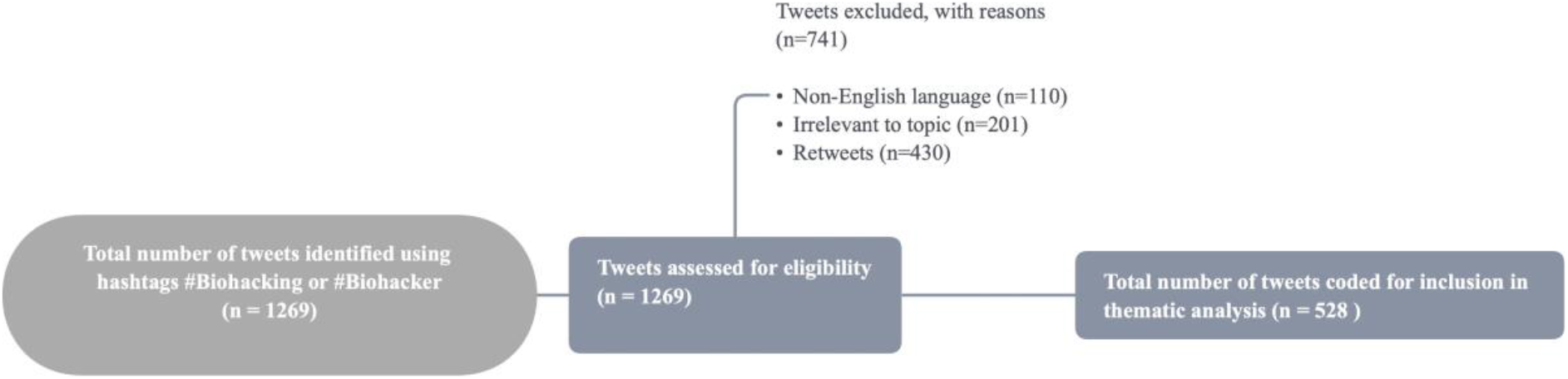
flow diagram of findings

As all collected tweets were publicly available at time of collection, this study was not subject to review by an institutional ethical review board [18,19].

### Thematic Analysis

Thematic analysis was employed to explore peoples’ discussions of biohacking on Twitter. An inductive approach was applied during the manual coding process, using NVivo V12 [20]. The questions asked during identification of themes were: “Is this tweet aimed at health improvement?” and “What is the key message in the tweet and what theme does this fall under, if not health?”.

Once coding was complete for all tweets that were deemed to be ‘on topic’, they were merged where deemed appropriate. As an example, ‘nootropics’ and ‘smart drugs’ were merged into a single code called ‘nootropics’. Thereafter, the codes were grouped into broader organising themes e.g., physical health and nutrition. This was an iterative process with progressive grouping of themes into more general parent themes. Initial coding and presentation of the data was done by QR. LCC and HMM contributed to interpretation of the coded data.

As nutrition formed a major theme of the analysis with many subtopics, a sunburst diagram was created to demonstrate the various nutrition-related topics discussed in a hierarchical and proportional representation (see Figure 3).

**Figure 3:**
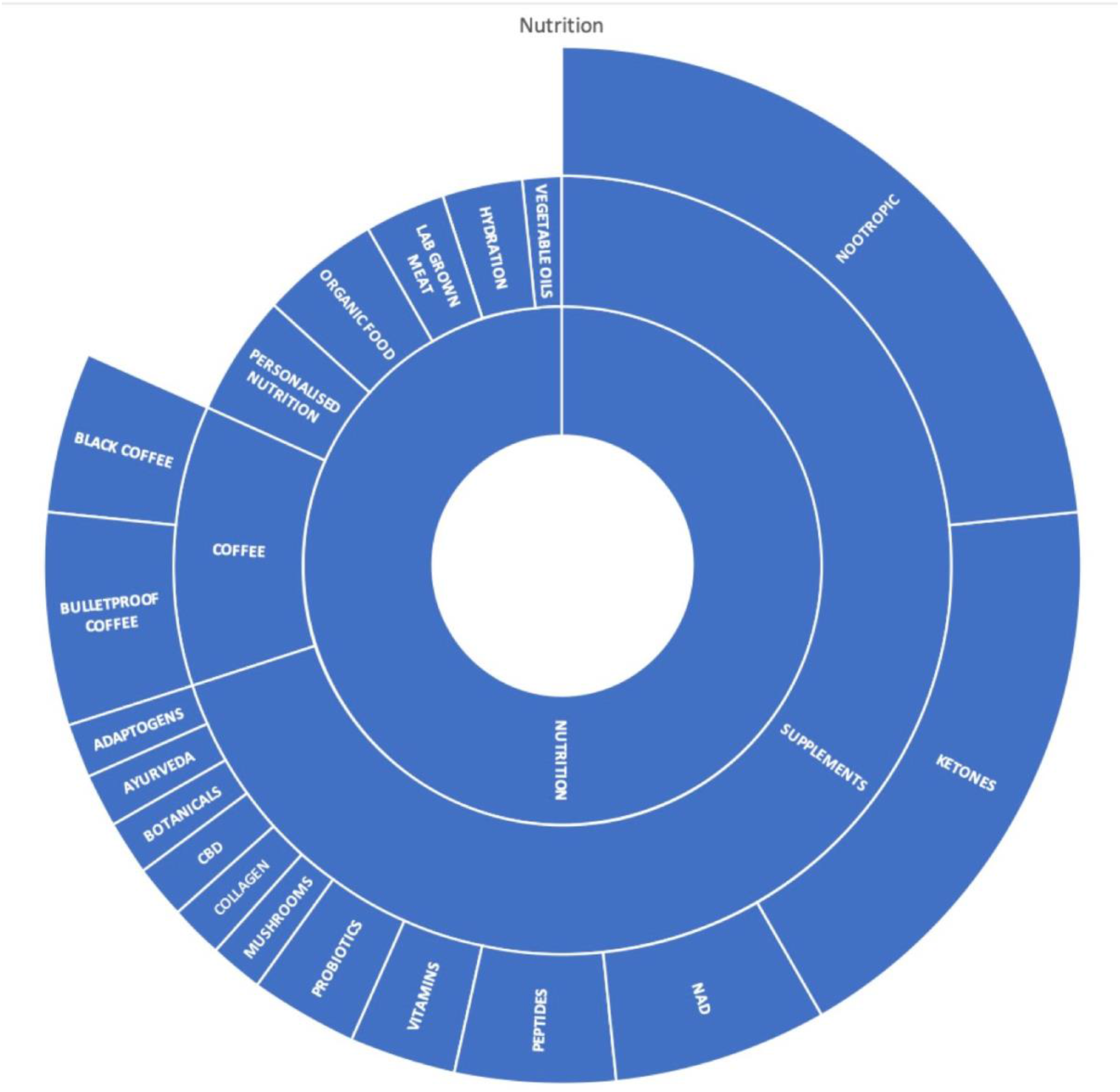
Sunburst diagram of topics discussed under the theme of nutrition. The diagram is hierarchical and proportional according to number of nodes.

## Findings

Of the total 1269 tweets identified using hashtags #biohacker and #biohacking, all were assessed for eligibility, with a total of 741 excluded. Among the excluded tweets, 110 were not in the English language, 201 were deemed to be non-sensical or irrelevant to biohacking and 430 were retweets. The remaining 528 tweets were included in the thematic analysis (see Figure 2).

Early in the coding process, it became apparent that most of the tweets were health centric, and content within each tweet belonged to a multitude of themes, often with an overarching theme of health. Nutrition comprised the largest theme under health and this contrasts to some of the definitions of biohacking in published literature where technology, wearables and embedded implants are commonly referenced [21]. Thematic analysis revealed substantial heterogeneity when it comes to what Twitter users perceive to constitute biohacking. The tweets ranged from basic lifestyle advice such as sleep and nutrition to more invasive suggestions such as subdermal chip implants. An example of a tweet about health is “The same things that kept you healthy for centuries still keep you healthy now! Sleep, Sunlight, Exercise, Real food”. However, a tweet by another user refers to a more invasive form of biohacking: “…small, passive, sensor-equipped injectable subdermal implants that allow users to check their vital parameters”.

The three major themes were physical health, psychology, and science. Within each theme there was a wide variation in topics (Table 1) and these are discussed in more detail.

**Table 1:** themes and topics identified, alphabetically organised

| **Physical health** | **Psychology** | **Science** |
| --- | --- | --- |
| Aging | Emotional Intelligence | Antibiotic resistance |
| Alternative medicine | Emotions | Astrology |
| Biomarker Testing | Focus | Biology |
| Brain | Gratefulness | Capacitance |
| Cardiovascular Health | Habits | COVID-19 Vaccine |
| Dental Health | Meditation | DIY Science |
| Detox | Mental health | Electromagnetic Frequency |
| Disease | Mind body spirit | Evidence Based Biohacking |
| Gut health | Mindfulness | Exclusion Zone Water |
| Hair | Mindset | Genetics |
| Healing | Monk | Heat Shock Protein |
| Healthcare | Productivity | Interstitial Pressure |
| Hormones & Neurotransmitters | Psychedelics | Metabolism |
| Immune System | Relaxation | Microbiome |
| Light Therapy | Resilience | Neuroscience |
| Nutrition | Self-development | Nuclear Respiratory Factor<br>(NRF) |
| Physical Fitness | Spirituality | Stem cells |
| Physical therapy modalities | Stress | Toxins |
| Precision Medicine | Trauma | Spiderbeer |
| Reproductive & Sexual Health | Work culture |  |
| Skin |  |  |
| Sleep |  |  |
| Wellness |  |  |

### Physical Health

The most popular topics discussed within the theme of physical health included physical fitness, nutrition, wellness, aging, hormones/neurotransmitters and sleep (Table 1).

#### Physical Fitness

Tweets on physical fitness included the topics of performance, exercise, and breathwork. Tweets about exercise referenced benefits of various modalities of exercise including weightlifting, cardiovascular exercise, CrossFit®, high intensity interval training (HIIT). Tweets on breathwork emphasised the many benefits of breathwork for general health, with much ambiguity as to its exact benefits. The other topics with minimal tweets discussed fitness, yoga, bone density, sport, and blood flow restriction training.

On the topic of physical fitness, one Twitter user tweeted:

When the legs lose their pep, what’s an aging distance runner to do?

1. Wallow in self-pity
2. Embark in #biohacking journey, using #IoT wearables & custom #MachineLearning models. Leveraging: @Azure @whoop @MyFitnessPal @GarminFitness

#### Nutrition

Tweets discussing nutrition included specific nutrients, supplements, and diets, as shown in Figure 3. Supplements and coffee comprised most of the discussion within nutrition.

Supplements discussed included nootropics, ketones, nicotinamide adenine dinucleotide (NAD), peptides and probiotics. The proposed benefits of supplements centred on improving cognition, mental clarity, and general physical wellbeing. Discussion on coffee was polarised with discussion about black coffee, as well as bulletproof coffee, with the latter often having butter or another fat source added to it [22]. Other tweets recommended a move towards use of personalised nutrition; lab grown meat; organic foods and avoidance of seed oils to maintain good health. A couple of tweets emphasised going back to fundamentals such as eating vegetables and ensuring adequate hydration status. One user tweeted “You should add broccoli to your diet. It provides you with multiple vitamins & minerals: Vitamin C, Zinc, Magnesium, Selenium, Sulforaphane, Indoles”. Another user tweeted “Hydration is often overlooked in terms of importance for health. Being dehydrated is one of the main things that can negatively impact testosterone production.” A few tweets conveyed the users’ perceptions of the importance of blood tests to determine nutrient status for personalising diet. Nutrition is a key topic of discussion relevant to biohacking.

#### Wellness

Wellness was used in the context of physical and mental health, although there was a lot of ambiguity in many of the tweets as to exactly what this means. Wellness was included under the topic of physical health as this was the predominant context within which this term was used. Those tweeting about biohacking and using ‘wellness’ often took a holistic approach, as demonstrated by the use of this word in the contexts within which it has been used. Using #wellness, one user tweeted “Your State Of Mind Is Everything”, and another quoted Dalai Lama “Happiness is not something ready made. It comes from your own actions. - Dalai Lama”. Many of the practices in this theme do not require the use of technology, drugs, or other substances.

#### Aging

Aging related discussions included methods of achieving longevity, supplements and lifestyle habits purported to slow aging. One user commented “You can’t control chronological age, you can, however, slow cellular aging.” One user commented that their telomere test suggested their biological age was less than their chronological age. The emphasis on aging was on living healthier and longer. One user tweeted “Testosterone is anti-aging for men”.

These practises involve a combination of technology, drugs, other chemicals and lifestyle changes.

#### Hormones and neurotransmitters

There was an emphasis on testosterone and cortisol amongst tweets discussing hormones, with dopamine being the main neurotransmitter discussed. Some of the tweets discussed methods to optimise testosterone such as minimising dehydration, reducing intake of ultraprocessed foods, and increasing intake of specific nutrients, and improving other general lifestyle habits.

Testosterone was mentioned in multiple tweets with an example shown below: “Optimized testosterone production means:

Lower body fat

More muscle mass

Stronger bones

Better circulation

Healthier heart.”

Cortisol was also mentioned with one user tweeting “Box breathing exercises are an excellent way for you to reduce stress levels & the stress hormone cortisol”.

#### Sleep

Discussions on sleep included methods of “biohacking the circadian rhythm”, use of blue light blocking glasses, sleep monitoring methods and general lifestyle changes to improve sleep quality. One user tweeted about a sleep tracking app: “Sleep Cycle App Review: A sleep quantification tool and precision alarm clock. An app that all real Biohackers need: Sleep Cycle alarm clock”.

#### Disease and pathology

Inflammation and insulin resistance formed the bulk of discussions about disease and pathology, with tips on methods of reducing inflammation and improving insulin resistance focussing on dietary and lifestyle changes. The other discussions were about pain management, autoimmune disease and one tweet was about the impact of dental health on the immune system and inflammation, as one user tweeted: “How Your Teeth Are Impacting Your Immune System, Sleep, Performance, Inflammation”

#### Physical Therapies

This theme included practices that were discussed in the context of improving health and included grounding, sun exposure, pulsed electromagnetic magnetic field therapy, biofeedback, massage, bioresonance therapy, ozone treatment, sauna, cold exposure, and hyperbaric oxygen therapy. Grounding is the practice of making direct skin contact with the ground [23]. The tweets referring to grounding suggested that this practice improves sleep and reduces inflammation. An example of such a tweet is “The Science of Grounding: How Earthing Improves Sleep and Reduces Inflammation”. Once again, some practices here such as massage, sun exposure and grounding involve no technology, drugs or other substances.

### Psychology

The discussions related to psychology were generally about improving mental health using a variety of techniques such as mindfulness, meditation, and self-development with a couple of hashtag references to #psychedelics. The two tweets that mention psychedelics as hashtags make no reference to any particular use other than as a means of biohacking. However, interest in psychedelics is growing as a form of therapy to treat a variety of mental health conditions including depression and post-traumatic stress disorder [24]. The full list of topics discussed can be seen in Table 1.

Some of the tweets suggested stressing less to improve other aspects of physical or metabolic health such as testosterone whilst others advised on physical lifestyle changes that could be made to improve mental health. An example is “drop the ultraprocessed food and mental health will improve” and another tweet suggested that increased muscle mass could reduce risk of depression.

The references to spirituality, Buddhism and Taoism suggest that various spiritual practices form part of some biohacking routines.

### Science

More esoteric topics were grouped under science where the tweets were focussed on scientific fields or specific scientific technologies, that were not explicitly related to any of the other themes. Discussion of genetics was the most popular topic within this theme, spanning nutrigenomics, genetic modification and Clustered Regularly Interspaced Short Palindromic Repeats (CRISPR) technology.

Other topics and keywords mentioned included biology, nuclear respiratory factor (NRF), synthetic biology, stem cells, heat shock proteins, metabolism, microbiome, spiderbeer and toxins. Spiderbeer refers to production of silk by inserting the silk gene from a spider into a plasmid [25]. There was one tweet that asked, “What the best place to start to learn about the latest (evidence based) #biohacking?”, raising an interesting point about the evidence base behind this movement.

There were several tweets referring to do it yourself (DIY) science, centred around the ease with which the COVID-19 virus and vaccines could be engineered out with traditional settings. An example is “Thanks to a technological revolution in genetic engineering, all the tools needed to create a #virus have become so cheap, simple, and readily available that any rogue scientist or college-age #biohacker can use them, creating an even greater threat.”

This tweet emphasised how technological advancements have significantly lowered the barriers to allow people to engage in bioengineering out with formal establishments.

#### Human Enhancement

Implants were a popular discussion topic for methods of human enhancement and included near field communication (NFC) subdermal chips, brain chips. One twitter user stated “Embrace it. It’s inevitable. We’re all going to slowly merge with the machine.”

## Discussion

### Implications

Despite increasing adoption of consumer health-related technologies and practices, published literature on biohacking remains sparse. The existing literature often refers to biohacking with practices such as genetic modification and subdermal chip implants [26,27]. However, this study highlights that this does not seem to be the main practice of those engaging in biohacking, with many practices requiring no technology, drugs or other substances. This suggests that practices deemed to be biohacking are likely to be much more prevalent than many would consider according to the definitions mentioned. To our knowledge, this study is the first to explore publicly available discussions on biohacking, and the themes and topics reveal that most people could be deemed to be engaging in biohacking practices, even if they do not identify as biohackers. This practice could be as simple as taking a dietary supplement or meditating. This study highlights the importance of ongoing research in this field of unregulated consumer health-related technologies, especially as access to and sharing of information and knowledge continues to increase with the use of social media and resources on the internet.

These findings raise the question as to whether the definition of biohacking needs to be refined to include a broader conceptualisation of practices, accounting for intention, early adoption and self-identity. A refined definition: “biohacking refers to *intentional* attempts *by early adopters* to improve the condition of their body and mind, often using technology, drugs, or other chemical substances such as hormones”.

Despite the perception among some people that biohacking must involve some form of technology, our findings show that this is not necessarily the case. This study demonstrates how all-encompassing the perception of biohacking is on Twitter, incorporating most aspects of basic human needs through to cutting edge technology [21]. For example, nutrition is a fundamental human need and has been practiced in various ways throughout human history and the act of eating is not technological in and of itself. However, what has changed is our understanding of nutrition and ability to share and access information online, enabled by advances in technology. These advances have allowed for better understanding of nutrients, their health benefits as well as allowing nutritional intake to be tracked [28]. More specifically, physical fitness and nutrition comprise a significant proportion of discussions on biohacking which does not align with definitions in published literature on biohacking. Yetisen refers to biohacking as “do-it-yourself citizen science merging body modification with technology” with the motivation of biohacking including “cyberkinetic exploration, personal data acquisition, and advocating for privacy right and open-source medicine”. Whilst the findings in this study include some of these topics, it expands well beyond to include more basic practices that require no technology or data acquisition such as sun exposure and dietary change [29]. Another paper refers to biohacking as “optimising one’s physiology through practices including intermittent fasting, elimination diets, targeted supplementation, and multiple cycles of androgenic anabolic steroids” [30]. The heterogeneity in what constitutes biohacking in published literature is apparent and this study advances our understanding of how biohackers conceptualise their practices and future research should build upon these findings.

With regards to technology, wearables can provide useful health related data including heart rate, blood pressure, oxygen saturation, heart rate variability and rhythm [31]. However, there is heterogeneity in the quality of hardware and algorithms of these wearables. Inaccurate data can be more harmful than no data, especially if this data is used for diagnostic and treatment purposes. Integration of these technologies are rapidly progressing through the Gartner Hype Cycle [31], meaning consumers and healthcare providers should remain mindful of the potential for harm to occur.

With access to and provision of healthcare being affected during the COVID-19 pandemic, many have turned to self-care practices accompanied by an increase in people seeking information on health, often on social media [32,33]. This has the potential to cause harm if people follow health related advice without seeking healthcare input, especially those with pre-existing health conditions. An example is someone with coronary heart disease attempting to follow an exercise regime without consulting with their healthcare provider.

The COVID-19 pandemic has catalysed the number of people seeking health related information online, and thereby increasing the risk of exposure to misinformation and potentially dangerous practices [33].

### Strengths

Some strengths of this study are that the information is in the public domain on Twitter where biohackers are sharing information and there was a large dataset. This affords researchers an opportunity to repeat this study in future to explore changes in biohacking practices as technology continues to develop.

### Limitations

At present, tweets are limited to 280 characters. This poses a challenge for collecting comprehensive data on discussions on specific topics. In this study, we found that many tweets used bulleted lists or a collection of many hashtags, posing an issue for clear interpretation. Although threads, whereby multiple tweets are published in a chain, are often used to overcome this, these can be difficult to collect and analyse as each tweet was identified independently by TAGS software and even then, each tweet in the thread may not always include the relevant hashtag to allow it to be identified as relevant.

Relevant tweets published by self-identifying biohackers that did not use the hashtags #biohacker or #biohacking will have been excluded during initial data collection, as well as those that were not in the English-language. The lack of demographic profile data restricted further analysis involving deeper sociological meaning and makes it difficult to exclude fictional tweets.

Unlike other large social media platforms, tweets are public by default, and the application programming interface (API) makes access to these tweets readily available. However, there are limitations to the number of queries that can be run, capturing only a proportion of tweets relevant to our topic. The number of tweets not captured remains difficult to quantify.

Furthermore, tweets from users who choose to make their account private will have been excluded from our analysis. A recent survey showed that 13% of US users keep their twitter accounts private [34].

Despite searching Twitter using the hashtags #biohacker and #biohacking, there was an abundance of tweets that were irrelevant to the topic being studied. This included tweets linking to other websites, referencing human rights abuse or using lists or words that were incomprehensible. This is due to the fact that hashtags are user-generated, making them usable within any context.

This is the first known study to analyse discussions on biohacking, which meant that an inductive approach to the collection of tweets and their analysis was required to understand how ‘biohackers’ conceptualise their practices in their own terms, rather than applying *a priori* definitions from medical or academic perspectives. This approach was used due to restrictions of data exploration and collection on the Twitter platform and novelty of this topic for which social media provides an ideal platform for biohackers to share practices.

Whilst being convenient for research purposes, this does highlight the ease with which misinformation can be propagated with easy access. The themes identified in this study will allow further research to be undertaken using a variety of keywords identified to find discussions of more focussed biohacking topics. However, the search did not include the hashtag ‘#biohack’, and this may have excluded some tweets that did not also include any of the hashtags used for data collection.

Sentiment analysis, whereby positive or negative attitude is identified and analysed, was not possible due to the lack of sentiment expressed in the tweets. This was, in part, due to the use of the broad search terms using hashtags #biohacker and #biohacking and 280-character limitation imposed by Twitter.

### Recommendations

The general recommendations that arise from this study are for increased awareness amongst healthcare professionals about biohacking practices to facilitate effective conversations and documentation as patients may not always offer this information without being asked. There may be lethal consequences as a result of some biohacking practices such as an interaction between an anticoagulant and dietary supplement increasing bleeding or thrombotic risk [35]. As this study demonstrates, taking dietary supplements forms a large part of biohacking practice and these may have interactions with prescribed medication, affect organ function, and cause toxicity [36]. Furthermore, with increasing consumer genetic and blood testing available to consumers, it is crucial that healthcare professionals are aware of the potential that patients may be self-diagnosing and self-treating in the context of biohacking and such information needs to be accounted for when discussing treatment options, risks, and benefits. This study highlights how vast biohacking practices can be and suggests that healthcare professionals should routinely ask patients about any self-directed health practices that they engage in as not all patients will identify as a biohacker or be aware of potential harms.

## Conclusion

The aim of this study was to explore what participants on Twitter (twitter.com) are discussing using the term ‘biohacking’, collecting tweets that included relevant hashtags (#biohacking and #biohacker) to identify the themes and topics discussed. This study has highlighted the heterogeneity and variation in what the public discussion of biohacking is. Twitter users using biohacking hashtags posted about many topics related to the enhancement of health, drugs, and technology, often assisted by data acquisition, with the greatest contribution related to physical fitness and nutrition. Health appears to be important for people discussing biohacking, with a particular emphasis on physical fitness and nutrition which do not always require technology, drugs or other substances. The implications of unprecedented access to scientific knowledge, wearable and embedded health-related devices are immense, and the emergence of the biohacking community should be better understood with a view to a unified definition of biohacking. These results demonstrate the ambiguity underlying biohacking and call for enrichment of scientific discussion surrounding the topic of biohacking, incorporating deeper sociological components such as factors influencing adoption, risks of biohacking practices and how healthcare professionals can assist in identifying and mitigating these.

Given the changes that the internet and technology has brought about in shaping human behaviour, big changes lie ahead and as we move towards an increasingly connected world with more affordable technology, it is imperative we are aware of the various movements being shaped. These changes can benefit the health of the population but also afford much opportunity for harm. This information should allow for public health and healthcare professionals to implement policy surrounding education, regulation, and safe use of such technologies as society inevitably continues to adopt technology at unprecedented rates.

## Data Availability

The raw data are not available due to their potentially identifiable nature.

## Author Biographies

### Qasim Rafiq

Qasim is a medical doctor and associate registered nutritionist whose formal training includes nutrition, lifestyle medicine and sexual health. He is currently working in General Practice within NHS Scotland.

### Lynsey Christie

Lynsey M. Christie, PhD, is a Lecturer in the School of Pharmacy and Life Sciences, at Robert Gordon University, Scotland. Working in different areas of applied health her research has spanned various topics but has mostly focused on the impact of different dietary components on cardiovascular disease development.

### Heather May Morgan

Heather is a multidisciplinary social scientist whose formal training spans law, French language, forensic medicine, philosophy, gender studies, social research, sociology, criminology and health services research. She is a Lecturer (Scholarship) within the University of Aberdeen’s [Postgraduate Education Group, Institute of Applied Health Sciences](https://www.abdn.ac.uk/iahs/research/postgraduate-education/index.php). She has substantive research interests in digital health and methodological expertise in designing, leading and delivering qualitative and mixed methods studies in applied health sciences. To date, she has individually or jointly secured research funding totalling more than £1.6m. She is lead author or co-author of over thirty peer-reviewed papers and two edited collections and is a public engagement with research prize-winner.

## References

[1] Cambridge University Press. Meaning of biohacking in English. n.d.; Available at: https://dictionary.cambridge.org/dictionary/english/biohacking. Accessed September/6, 2021.

[2] Greiwe J, Nyenhuis SM. Wearable Technology and How This Can Be Implemented into Clinical Practice. Current allergy and asthma reports 2020;20(8):36.

[3] Berman T. The Smart Ring: From the 17th Century Wearable Abacus to Today. 2017; Available at: https://interestingengineering.com/smart-ring-17th-century-wearable-abacus-today. Accessed 08/22, 2021.

[4] Isakadze N, Martin SS. How useful is the smartwatch ECG? Trends Cardiovasc.Med. 2020;30(7):442–8.

[5] Reed MJ, Grubb NR, Lang CC, O’Brien R, Simpson K, Padarenga M, et al. Multi-centre Randomised Controlled Trial of a Smartphone-based Event Recorder Alongside Standard Care Versus Standard Care for Patients Presenting to the Emergency Department with Palpitations and Pre-syncope: The IPED (Investigation of Palpitations in the ED) study. EClinicalMedicine 2019;8:37–46.

[6] Internet world stats. Internet growth statistics. n.d.; Available at: https://www.internetworldstats.com/emarketing.htm. Accessed June/16, 2021.

[7] Gerbis N. How biohacking works. n.d.; Available at: https://science.howstuffworks.com/innovation/scientific-experiments/biohacking.htm. Accessed January/12, 2021.

[8] Vailshery L. Number of connected wearable devices worldwide from 2016 to 2022. 2021; Available at: https://www.statista.com/statistics/487291/global-connected-wearable-devices/. Accessed June/, 2021.

[9] Bujnowska-Fedak M, Węgierek P. The Impact of Online Health Information on Patient Health Behaviours and Making Decisions Concerning Health. International journal of environmental research and public health 2020 January 31;17(3):880.

[10] Ireland T. Do It Yourself. n.d.; Available at: https://www.rsb.org.uk/biologist-features/the-unlikely-labs. Accessed 08/29, 2021.

[11] Hesketh J. Personalised nutrition: how far has nutrigenomics progressed? Eur.J.Clin.Nutr. 2013 May 01;67(5):430–5.

[12] Vicente AM, Ballensiefen W, Jönsson J. How personalised medicine will transform healthcare by 2030: the ICPerMed vision. Journal of Translational Medicine 2020 April 28;18(1):180.

[13] Mathur S, Sutton J. Personalized medicine could transform healthcare. Biomedical reports 2017;7(1):3–5.

[14] Meskó B, Drobni Z, Bényei É, Gergely B, Győrffy Z. Digital health is a cultural transformation of traditional healthcare. mHealth 2017 September 14;3:38.

[15] Sinnenberg L, Buttenheim AM, Padrez K, Mancheno C, Ungar L, Merchant RM. Twitter as a Tool for Health Research: A Systematic Review. Am.J.Public Health 2017;107(1):e1–8.

[16] Twitter. Counting characters. n.d.; Available at: https://developer.twitter.com/en/docs/counting-characters. Accessed October 14, 2021.

[17] Hawksey M. Twitter archiving Google spreadsheet TAGS v6.1. 2014; Available at: https://tags.hawksey.info/get-tags/. Accessed August, 2020.

[18] Skea ZC, Entwistle VA, Watt I, Russell E. ‘Avoiding harm to others’ considerations in relation to parental measles, mumps and rubella (MMR) vaccination discussions - an analysis of an online chat forum. Soc.Sci.Med. 2008 November 01;67(9):1382–90.

[19] Shah F, Bhattacharya S, Lamont K, Morgan HM. “Your womb, your choice!” Making an informed decision regarding the timing of pregnancy following miscarriage. medRxiv 2020 January 01:2020.09.10.20191858.

[20] QSR International Pty Ltd. NVivo (released in March 2020). 2020;.

[21] Gangadharbatla H. Biohacking: An exploratory study to understand the factors influencing the adoption of embedded technologies within the human body. Heliyon 2020;6(5):e03931.

[22] Sissons C. What are the health benefits of bulletproof coffee? 2018; Available at: https://www.medicalnewstoday.com/articles/323253#health-diet-and-lifestyle-benefits. Accessed June/01, 2021.

[23] Menigoz W, Latz TT, Ely RA, Kamei C, Melvin G, Sinatra D. Integrative and lifestyle medicine strategies should include Earthing (grounding): Review of research evidence and clinical observations. EXPLORE 2020 May 01;16(3):152–60.

[24] Tupper KW, Wood E, Yensen R, Johnson MW. Psychedelic medicine: a re-emerging therapeutic paradigm. CMAJ : Canadian Medical Association journal = journal de l’Association medicale canadienne 2015 October 06;187(14):1054–9.

[25] The Thought Emporium. Spider Beer Overview. n.d.; Available at: https://www.thethoughtemporium.com/spiderbeer. Accessed September/02.

[26] Shinde S, Meller-Herbert O. Biohacking. Anaesthesia 2017 July 01;72(7):909.

[27] Nash DB. Beware biohacking? Biotechnol.Healthc. 2010 January 01;7(1):7.

[28] Ferrara G, Kim J, Lin S, Hua J, Seto E. A Focused Review of Smartphone Diet-Tracking Apps: Usability, Functionality, Coherence With Behavior Change Theory, and Comparative Validity of Nutrient Intake and Energy Estimates. JMIR mHealth and uHealth 2019 May 17;7(5):e9232.

[29] Yetisen AK. Biohacking. Trends Biotechnol. 2018 August 01;36(8):744–7.

[30] Nagata JM, Brown TA, Lavender JM, Murray SB. Emerging trends in eating disorders among adolescent boys: muscles, macronutrients, and biohacking. Lancet Child.Adolesc.Health. 2019 July 01;3(7):444–5.

[31] Bayoumy K, Gaber M, Elshafeey A, Mhaimeed O, Dineen EH, Marvel FA, et al. Smart wearable devices in cardiovascular care: where we are and how to move forward. Nature reviews.Cardiology 2021;18(8):581–99.

[32] Fiske A, Schneider A, McLennan S, Karapetyan S, Buyx A. Impact of COVID-19 on patient health and self-care practices: a mixed-methods survey with German patients. BMJ Open 2021 September 01;11(9):e051167.

[33] Neely S, Eldredge C, Sanders R. Health Information Seeking Behaviors on Social Media During the COVID-19 Pandemic Among American Social Networking Site Users: Survey Study. J.Med.Internet Res. 2021 June 11;23(6):e29802.

[34] Remy E. How public and private Twitter users in the U.S. compare — and why it might matter for your research. 2019; Available at: https://medium.com/pew-research-center-decoded/how-public-and-private-twitter-users-in-the-u-s-d536ce2a41b3. Accessed 22 May, 2021.

[35] Tarn DM, Barrientos M, Wang AY, Ramaprasad A, Fang MC, Schwartz JB. Prevalence and Knowledge of Potential Interactions Between Over-the-Counter Products and Apixaban. J.Am.Geriatr.Soc. 2020;68(1):155–62.

[36] Peng CC, Glassman PA, Trilli LE, Hayes-Hunter J, Good CB. Incidence and Severity of Potential Drug–Dietary Supplement Interactions in Primary Care Patients: An Exploratory Study of 2 Outpatient Practices. Arch.Intern.Med. 2004;164(6):630–6.

